# Beyond COVID-19 Deaths: Cause-Specific Analysis of Excess Mortality in Russia

**DOI:** 10.64898/2026.03.23.26349084

**Authors:** Ekaterina Degtiareva, Sergey Timonin, Andrea Tilstra, José Manuel Aburto

## Abstract

During the COVID-19 pandemic, European mortality exhibited a marked East-West divide in both timing and magnitude, echoing longstanding longevity disparities in this region. Russia sits on the Eastern side: early restrictions were short-lived, and vaccine uptake remained low amid historically limited trust in government and science. Using weekly national and monthly regional mortality data disaggregated by age, sex, and cause of death, we estimated excess mortality from March 2020 to December 2021 using generalised additive models. We identify two major mortality peaks (late 2020-early 2021 and late 2021) and estimate 1,044,914 excess deaths, well above the 595,815 officially registered COVID-19 deaths. Non-COVID-19 excess was larger during the first peak, especially at ages 15–44. Cardiovascular diseases accounted for roughly 60% of the non-COVID-19 excess and we find no evidence of excess mortality from cancer or external causes. Among women, excess deaths were concentrated at older ages, whereas among men they clustered at working and older working ages, only partly reflecting differences in age structure. The highest excess mortality was found in the most populous regions, particularly the Central European and Volga parts. Temporal and spatial inconsistencies in cause-of-death coding may obscure indirect mortality burden and hinder the associated policy response.

**Highlights:**

- Russia had 1,044,914 excess deaths in 2020–21, about twice official COVID-19 deaths.
- These discrepancies varied over time and across regions.
- Cardiovascular deaths drove most non-COVID excess mortality.
- We find no evidence of excess mortality from external causes of death.
- Autopsy-based COVID-19 assignment may have increased misclassification

## Introduction

A pronounced East-West divide deepened in Europe during the COVID-19 pandemic: countries east of the former Iron Curtain — Central and Eastern Europe including the post-Soviet states — generally recorded higher excess mortality than Western Europe (Aburto, Schöley, Kashnitsky, Zhang, et al., 2022a; Islam et al., 2021; V. M. Shkolnikov et al., 2023). They also witnessed lower vaccine uptake and less sustained non-pharmaceutical interventions (NPIs), patterns often linked to institutional capacity and lower levels of public trust (Bonnet et al., 2024; V. M. Shkolnikov et al., 2023). Russia sits firmly on this Eastern side of the divide, with one of the largest absolute excess mortality burdens and the steepest life expectancy losses in Europe and worldwide (Aburto, Schöley, Kashnitsky, & Kashyap, 2022; V. M. Shkolnikov et al., 2023; Wang et al., 2022). After a brief nationwide shutdown in spring-early summer 2020, restrictions were eased and subsequent NPIs were heterogeneous across regions. Like its other Eastern Europe counterparts, vaccination uptake in Russia also lagged despite domestic vaccine availability, reflecting widespread hesitancy and limited institutional trust (Lamberova & Sonin, 2023; Peshkovskaya & Galkin, 2023; Scherbov et al., 2022).

Excess mortality during the COVID-19 pandemic, an indicator used to address comparability and under-ascertainment of causes of deaths, has been estimated to be one of the highest for Russia globally (Beaney et al., 2020; Wang et al., 2022). A recent study suggests more than one million excess deaths was observed in Russia in 2020-2021 (Kobak, 2021), yet evidence remains limited on how these losses are distributed across causes, regions, or demographic groups (Makarova et al., 2021; Timonin et al., 2022). The substantial gap between COVID-19 deaths and excess mortality raises concerns about cause-of-death classification and reporting practices. In Russia, official COVID-19 mortality has relied on comparatively restrictive certification criteria (often autopsy-based), which may shift deaths involving SARS-CoV-2 toward other underlying causes (Dyer, 2021; Ivanova & Tsvetkova, 2020). Corresponding life expectancy losses were also severe: between 2019 and 2020, life expectancy fell by 1.68 years for men and 1.80 years for women, with further declines in 2021 (Aburto, Schöley, Kashnitsky, & Kashyap, 2022; Aburto, Schöley, Kashnitsky, Zhang, et al., 2022b; Islam et al., 2021). Recent comparative work suggests even larger cumulative losses by 2021 (Shkolnikov et al., 2023). These features make Russia a consequential case for how surveillance, death certification practices, and policy environments shape observed pandemic mortality.

This study uses official mortality data from Russia’s Federal State Statistics Service to estimate excess deaths from March 2020 to December 2021 along two dimensions: (1) national excess deaths disaggregated by age group, sex and cause of deaths; and (2) regional variation in all-cause excess mortality and officially registered COVID-19 mortality. We employ compositionally robust methods that capture seasonality and long-term trends while preserving add-up constraints across causes. Deaths are grouped into COVID-19, ischaemic heart disease (IHD), other cardiovascular diseases (CVD), respiratory diseases, cancers, external causes, substance-related causes, suicides, and a residual category, with analyses focused on ages 15-44, 45-74, and 75+. We identified two major surges (late 2020-early 2021 and late 2021), with total excess deaths approximately twice the number of officially registered COVID-19 deaths. Among non-COVID causes, ischaemic heart disease and other cardiovascular disease together account for more than half of the excess, suggesting a combination of potential misclassification, especially during the first major peak, and indirect effects of the pandemic. Spatially, excess mortality shows a distinct regional pattern, concentrated in central and southern regions rather than following the pre-pandemic north-east gradient.

## Background

### The COVID-19 Pandemic and Mortality

The COVID-19 pandemic, caused by the coronavirus SARS-CoV-2, began in late 2019 and rapidly evolved into a global crisis with profound health, social and economic consequences (Alizadeh et al., 2023; Chetty et al., 2024; Lopez-Leon et al., 2021; Nicola et al., 2020). Governments adopted various combinations of non-pharmaceutical interventions (NPIs) — including but not limited to stay-at-home orders, school and business closures, mask mandates and mobility restrictions — to limit transmission of the virus and reduce mortality (Fadlallah et al., 2023). These measures, together with behavioural responses and health-system strain, produced collateral effects on routine care, chronic disease management, mental health, and substance use (Avena et al., 2021; Buera et al., 2021; Clair et al., 2021; Fekadu et al., 2021; Kontoangelos et al., 2020; Moynihan et al., 2021).

Global assessments using excess mortality and life-expectancy metrics show large, heterogeneous impacts of the COVID-19 pandemic across countries and over time (Aburto, Schöley, Kashnitsky, Zhang, et al., 2022b; Karlinsky & Kobak, 2021; Wang et al., 2022). Many studies document severe life-expectancy losses and elevated excess mortality during 2020-2021, with important temporal variation (Islam et al., 2021; V. M. Shkolnikov et al., 2023). Excess mortality — the difference between observed and expected deaths — became the preferred metric for measuring overall pandemic mortality burden because it captures both direct (COVID-19) and indirect (i.e. attributed to healthcare disruption, behavioural, environmental and other factors) effects and is robust to differences in cause-of-death coding across settings (Beaney et al., 2020). However, aggregate excess counts alone do not reveal which causes accumulated the indirect burden, nor do they identify likely pathways of misclassification. Cause-specific decomposition adds leverage in two ways. First, it can identify deaths that may have been misclassified (for example, COVID-19 fatalities assigned to ischaemic heart disease or other chronic diseases) by revealing unexpected rises in specific categories (Levitt et al., 2022; Msemburi et al., 2023). Second, it documents indirect effects (e.g., interruptions in cardiovascular care or delayed treatment for chronic conditions) that would not be captured through COVID-19 certification alone (Chen et al., 2023; Degtiareva et al., 2024; Wadhera et al., 2021). Cross-national cause-specific work is still limited (Polizzi et al., 2025), and there are substantial methodological challenges in comparing cause patterns across systems with different certification practices.

### The East-West Divide in Europe

A pronounced East-West divide in pandemic mortality emerged across Europe: many Central and Eastern European countries experienced larger excess mortality and greater life-expectancy losses than most Western European countries, particularly during late 2020 and again in late 2021 (Aburto et al., 2021; Islam et al., 2021; Wang et al., 2022). By the end of 2021, cumulative life-expectancy losses in several Eastern European countries exceeded two to three years, compared with losses typically below one year in much of Western Europe, although with notable heterogeneity within both regions (Aburto, Schöley, Kashnitsky, Zhang, et al., 2022b; Schöley et al., 2022). These differences are shown to have been associated with vaccine adherence, shorter or less sustained NPIs, and lower institutional trust (Demertzis & Klironomos, 2026; Sobczak & Pawliczak, 2022; Yaddanapudi et al., 2023). Differences in health-system capacity and baseline mortality profiles, particularly the higher burden of cardiovascular disease in parts of Eastern Europe, may have intensified both direct COVID-19 mortality and indirect deaths during periods of system strain (Hajdu et al., 2024; Shkolnikov et al., 2023). Together, these patterns motivate a closer examination of large Eastern European systems where policies, risk-taking behaviour and healthcare system performance systematically differ from those in the West.

### Russia Before the Pandemic

Russia entered the pandemic with a distinctive mortality baseline. Mortality from cardiovascular diseases, external and alcohol-related causes was much higher compared to Western and many Central European countries, despite marked improvements between 2003 and 2019 (Grigoriev et al., 2014; Nikoloski et al., 2023; V. Shkolnikov et al., 2013; Vishnevsky et al., 2017). Cardiovascular disease accounted for roughly one million deaths per year - about 46% of all registered deaths - placing Russia among the higher end of age-standardised CVD mortality internationally^1^ (Matskeplishvili & Kontsevaya, 2021). Although alcohol consumption fell markedly after 2003 following a series of anti-alcohol measures (Grigoriev & Andreev, 2015), associated with gains in life expectancy, hazardous drinking remained a major driver of working-age mortality and an important contributor to CVD risk (Danilova et al., 2020; Zamyatnina et al., 2025).

The pre-pandemic sex gap in life expectancy was large — almost 10 years in 2021 (men 65.6; women 74.3), reflecting the much higher burden of CVD, alcohol-related, and external-cause mortality among men^2^. This mortality pattern shapes the age–sex structure of the population: at ages 75-79 the male-to-female ratio is 0.47 and declines further at older ages (World Population Prospects, 2024). Such demographic structure influences the distribution of pandemic deaths by sex and age and must be taken into account when interpreting aggregate outcomes.

Spatial inequalities in mortality are also substantial in Russia, with a persistent southwest-northeast mortality gradient and large regional and sub-regional differences in health-system capacity(Shchur & Timonin, 2021; Timonin et al., 2020). Although Russia maintains extensive healthcare infrastructure - including one of the highest numbers of hospital beds per capita globally - resources are distributed unevenly across regions (King & Dudina, 2021). In 2018, Russia had approximately 71.1 hospital beds and 37.4 physicians per 10,000 population, yet funding levels and service quality varied substantially between major urban centres and smaller towns or rural areas (Aleksandrova et al., 2019). These disparities reflect long-standing socio-economic differences, environmental conditions and cause-of-death profiles (notably alcohol- and external-cause mortality at working ages), overlaid on a governance model that is formally centralised but unevenly implemented across regions, resulting in substantial variation in resources and service quality (Nikoloski et al., 2024; Shishkin et al., 2022). Regional inequality in baseline health and in the ability to deliver care provides a plausible source of heterogeneity in pandemic impacts and in the indirect mortality effects.

### Russian Pandemic Responses, Vaccination and Death Certification Practices

Pandemic policies in Russia reflected and, in some respects, intensified the pre-existing conditions. Russia implemented a brief nationwide lockdown in spring-early summer 2020, including paid “non-working days,” school closures, and mobility restrictions, but these measures were lifted relatively quickly. Responsibility for subsequent control was largely delegated to regional authorities, producing heterogeneous and often weakly enforced local measures, including travel restrictions, self-isolation requirements, remote working and studying arrangements, and the introduction of QR-code health passes to regulate access to public facilities and transport (Blackburn et al., 2023; Policy Responses to COVID-19 in the Russian Federation, 2020). Stringency fluctuated across pandemic waves, tightening during periods of rising infections in late 2020 and again in mid-to-late 2021, but overall remained moderate relative to many Western European countries (Stronski, 2021).

Although the main domestically developed vaccine (Sputnik V) was authorised in December 2020, followed by two other vaccines (EpiVacCorona and CoviVac), vaccine uptake remained low: by mid-October 2021 only about 36% of the population had received at least one dose, despite domestic availability (Burki, 2020; Lazarus et al., 2021; Roshchina et al., 2022; Scherbov et al., 2022). Surveys point to widespread vaccine hesitancy and distrust in official information as key barriers (Roshchina et al., 2022). In response to persistently low coverage, regional authorities launched information campaigns, set ambitious vaccination targets, and in some cases introduced mandatory vaccination requirements for certain occupational groups, particularly in healthcare and the public sector. Yet these targets remained unachieved.

Data collection and reporting of COVID-19 cases and deaths in Russia have raised substantial concern, both domestically and internationally (Dyer, 2020). This concern relates specifically to cause-of-death certification practices: despite national and international guidelines and extensive reliance on autopsies, official COVID-19 deaths were compiled under comparatively narrow criteria, resulting in large discrepancies between registered COVID-19 deaths and overall excess mortality (Scherbov et al., 2022). While part of this gap may reflect indirect effects of the pandemic (e.g., deaths linked to disrupted access to care and health-system strain) it is also likely to be related to misclassification, especially during the early stages of the pandemic and in certain regions (Ivanova & Tsvetkova, 2020). In practice, available estimates suggest more than one million excess deaths in Russia in 2020–2021, compared with roughly 0.6 million deaths officially assigned as due to COVID-19 (Scherbov et al., 2022). Consistent with this discrepancy, excess mortality rate estimates place Russia among the hardest-hit countries in Europe, with approximately 375 excess deaths per 100,000 population in 2020 (Wang et al., 2022).

### Remaining Knowledge Gaps

Despite existing evidence on overall excess mortality in Russia, important gaps remain. Most existing studies have focused on cumulative excess deaths over broad periods, offering limited insight into how the composition of excess mortality evolved across successive pandemic waves. Systematic cause-specific analyses that track temporal changes in mortality patterns are scarce, constraining the ability to distinguish direct COVID-19 mortality from indirect effects and potential misclassification in cause-of-death coding. Evidence is also limited on how these patterns differed by age and sex. Finally, while large regional disparities in both mortality impact and reporting practices are widely acknowledged, they have not been examined in a unified framework that links temporal dynamics, cause composition and spatial heterogeneity.

This study addresses these gaps by combining weekly national mortality data disaggregated by age, sex and cause of death with monthly regional totals for January 2015-December 2021, and by applying a compositionally coherent modelling strategy to estimate and decompose excess mortality for March 2020-December 2021. This integrated approach allows us to characterise the timing and composition of excess mortality across pandemic waves, to describe age- and sex-specific patterns, and to document regional heterogeneity in both overall excess mortality and discrepancies between excess deaths and officially registered COVID-19 deaths.

## Methods

### Data

#### National cause-of-death (weekly)

We used official, de-identified death records to derive weekly mortality counts for the entire territory of Russia, disaggregated by age group, sex, and cause of death for the period January 2015–December 2021. Causes are coded according to ICD-10^3^ and grouped into nine categories relevant from clinical and public health perspective: COVID-19, ischemic heart disease (IHD), other cardiovascular diseases (CVD), respiratory, cancer, substance-related, suicides, other external causes, and residual. We distinguish IHD from other CVD given its central role in Russia’s mortality profile and concerns that some pandemic-period deaths may have been misclassified as IHD (Roth et al., 2022). Ages are aggregated to 15-44, 45-74, and 75+ to stabilise weekly estimation in a compositional framework (finer splits produced convergence problems due to sparsity).

#### Regional all-cause (monthly)

Our regional analysis is based on monthly mortality data covering January 2015–December 2022. At this level, we distinguish only two cause-of-death categories: COVID-19 and all other causes combined. We estimate crude death rates per 100,000 population using a fixed denominator equal to the mean regional population in 2020–2021, which enhances comparability across regions of different sizes.

### Statistical Analysis and Modelling

#### Modelling Framework

We estimate expected deaths with a two-part strategy that combines generalised additive models (GAMs) (Hastie & Tibshirani, 1986) for totals with compositional (CoDa-transformed) GAMs for cause or regional shares (Bergeron-Boucher et al., 2017; Kjærgaard et al., 2019; Oeppen, 2008).

Let 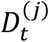 denote deaths for cause/region (j) at time (t) (week for national analyses; month for regions), λ_*t*_ the expected total deaths, and 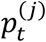 the expected share for cause (j) with 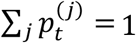. We impose coherence by modelling (1):

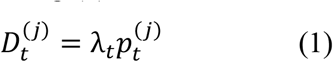

For totals, we fit Poisson GAMs with log link (2),

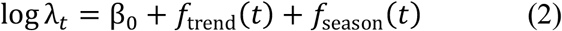

Where *f*_trend_(⋅) is a smooth long-run trend and *f*_season_(⋅) is a seasonal cyclic smooth. For composition, we transform the vector of shares *p*_*t*_ using the centered-log-ratio (clr) and fit GAMs to each clr-coordinate with the same trend and cyclic seasonality. Predictions are back-transformed to the simplex to obtain 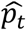, and coherent cause-specific expectations follow as 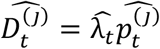. Within this clr step, uncertainty is propagated via a parametric bootstrap that draws from the approximate sampling distribution of GAM coefficients for both totals and clr-components, back-transforms to compositions, and combines with simulated totals to yield 1,000 predictive realisations and corresponding 95% intervals.

GAMs are not the only way to estimate expected deaths, so we benchmarked 4 most common in the excess mortality research models to identify the best fit. GAMs achieved the lowest RMSE and MAE for both weekly and monthly series^4^.

#### National analysis (Weekly data, Detailed causes of death)

Equations (1)-(2) are fit to weekly deaths by age-sex-cause using January 2015-February 2020 as the training period (baseline), with testing period predictions for March 2020-December 2021. Because expected COVID-19 deaths are identically zero in the training period, COVID-19 cannot enter the composition; by construction, COVID-coded deaths are absorbed into the model’s residual. For presentation (e.g., Figure 3), we subtract the officially coded COVID-19 series from this residual to display COVID and non-COVID residual separately. This does not affect excess values, as the expected COVID-19 deaths are zero.

#### Regional analysis (Monthly data, COVID-19 and all other deaths combined)

We apply the same framework with monthly seasonality to regional totals over January 2015-December 2022. Components *j* index **regions** (no cause breakdown). Excess is computed per region and summarised as rates per 100,000 using the fixed 2020-2021 average population. We compare total excess with officially registered COVID-19 deaths and report the residual (total excess minus COVID-registered), noting that differences reflect a mix of misclassification and indirect effects.

#### Model Fit and Excess Calculation

Models are fit to the pre-pandemic baseline (January 2015-February 2020) and used to generate predicted expectations for March 2020-December 2021. Excess deaths are defined as *observed* minus *expected* at the corresponding aggregation, and we compare these to officially assigned COVID-19 deaths, paying particular attention to alignment with IHD, respiratory, and other major categories.

All analyses are implemented in R 4.4.1 with standard CoDa utilities; reproducible code and an R implementation of the compositional routine (XCOD by Jonas Schöley, 2024) are provided in the Supplementary Material.

## Results

Overall, we estimated 1,044,914 excess deaths (95% CI: 1,012,626 to 1,076,741) across all causes and age–sex groups between March 2020 and December 2021 (Table 1). In contrast, the official COVID-19 death count over the same period was 595,815, accounting for approximately 57% of total excess mortality. Weekly excess deaths rose in two large waves (Figure 1): the first during winter 2020-2021 and the second in autumn 2021. While the official COVID-19 series closely tracks the timing of these waves, it consistently falls well below the all-cause excess. Sex-specific patterns differ in both timing and magnitude. Among women, the later period produced the single highest weekly excess, peaking in late October–November 2021, with a gap of roughly 5,300 deaths between total excess and officially registered COVID-19 deaths at the peak. Among men, excess mortality rose more sharply toward the end of 2020 and reached a lower peak in the second wave than among women. Nevertheless, COVID-19– coded deaths captured only 56% of cumulative male excess (260,512 COVID-19 deaths versus 464,360 excess deaths; Table 1). Early-pandemic weeks show little or no excess mortality for either sex, followed by a rapid increase toward the end of 2020, a partial return toward baseline, and a large resurgence in mid-to-late 2021. The latter coincides temporally with the summer 2021 heatwave, which may have contributed to excess mortality during this period.

**Table 1.**
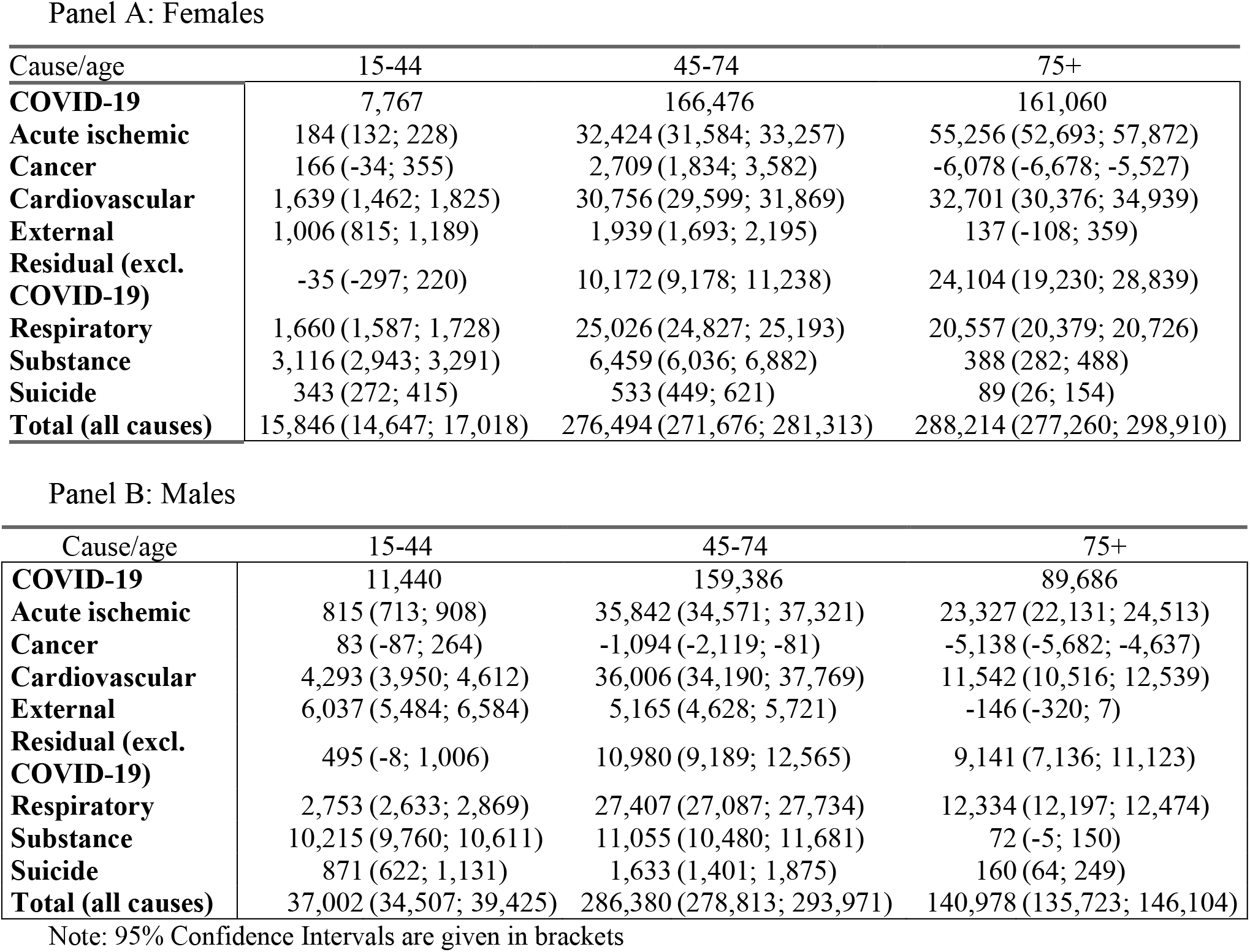
Total Cumulative Excess Deaths by Sex, Age and Cause. Russia, Mar 2020-Dec 2021.

Panel A: Females
| Cause/age | 15-44 | 45-74 | 75+ |
| --- | --- | --- | --- |
| **COVID-19** | 7,767 | 166,476 | 161,060 |
| **Acute ischemic** | 184 (132; 228) | 32,424 (31,584; 33,257) | 55,256 (52,693; 57,872) |
| **Cancer** | 166 (-34; 355) | 2,709 (1,834; 3,582) | -6,078 (-6,678; -5,527) |
| **Cardiovascular** | 1,639 (1,462; 1,825) | 30,756 (29,599; 31,869) | 32,701 (30,376; 34,939) |
| **External** | 1,006 (815; 1,189) | 1,939 (1,693; 2,195) | 137 (-108; 359) |
| **Residual (excl. COVID-19)** | -35 (-297; 220) | 10,172 (9,178; 11,238) | 24,104 (19,230; 28,839) |
| **Respiratory** | 1,660 (1,587; 1,728) | 25,026 (24,827; 25,193) | 20,557 (20,379; 20,726) |
| **Substance** | 3,116 (2,943; 3,291) | 6,459 (6,036; 6,882) | 388 (282; 488) |
| **Suicide** | 343 (272; 415) | 533 (449; 621) | 89 (26; 154) |
| **Total (all causes)** | 15,846 (14,647; 17,018) | 276,494 (271,676; 281,313) | 288,214 (277,260; 298,910) |

Panel B: Males
| Cause/age | 15-44 | 45-74 | 75+ |
| --- | --- | --- | --- |
| **COVID-19** | 11,440 | 159,386 | 89,686 |
| **Acute ischemic** | 815 (713; 908) | 35,842 (34,571; 37,321) | 23,327 (22,131; 24,513) |
| **Cancer** | 83 (-87; 264) | -1,094 (-2,119; -81) | -5,138 (-5,682; -4,637) |
| **Cardiovascular** | 4,293 (3,950; 4,612) | 36,006 (34,190; 37,769) | 11,542 (10,516; 12,539) |
| **External** | 6,037 (5,484; 6,584) | 5,165 (4,628; 5,721) | -146 (-320; 7) |
| **Residual (excl. COVID-19)** | 495 (-8; 1,006) | 10,980 (9,189; 12,565) | 9,141 (7,136; 11,123) |
| **Respiratory** | 2,753 (2,633; 2,869) | 27,407 (27,087; 27,734) | 12,334 (12,197; 12,474) |
| **Substance** | 10,215 (9,760; 10,611) | 11,055 (10,480; 11,681) | 72 (-5; 150) |
| **Suicide** | 871 (622; 1,131) | 1,633 (1,401; 1,875) | 160 (64; 249) |
| **Total (all causes)** | 37,002 (34,507; 39,425) | 286,380 (278,813; 293,971) | 140,978 (135,723; 146,104) |

Note: 95% Confidence Intervals are given in brackets

**Figure 1:**
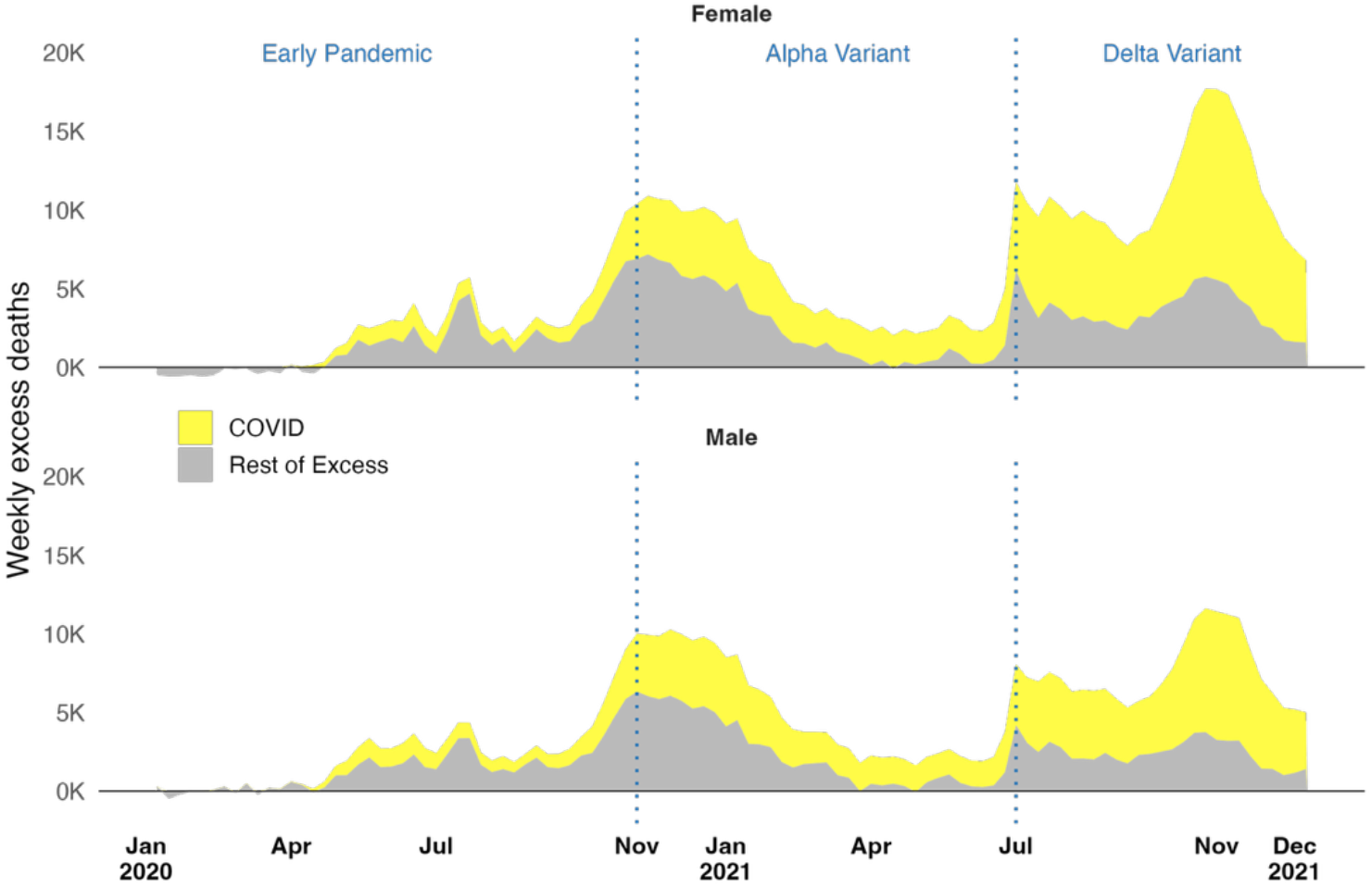
Weekly Excess Deaths by Sex, Russia, First Week of 2020 to Last Week of 2021.

Figure 2 decomposes the shortfall between excess deaths and official COVID-19 deaths. We plot only the pandemic period here, with observed deaths shown as dots and expected deaths as lines^5^. After the onset of the pandemic, the observed deaths diverged rapidly from the expected levels in both sexes. COVID-19-coded deaths are absorbed in the Residual category, which therefore rise steeply in both men and women. Increases in cardiovascular mortality— both IHD and other CVD—are evident above pre-pandemic baselines for men and women and they match the Residual dynamics, consistent with potential misclassification. Respiratory mortality shows pronounced winter pulses aligned in the same period and a smaller rise late in 2021. Cancer remains flat or slightly below expected levels for most of the period, contributing modest negative excess. Suicides show little deviation from trend (a slight decline for women and a small early-2020 uptick for men followed by stabilisation). Substance-related deaths increase notably for men beginning in mid-2020 and remain elevated thereafter, while other external causes move close to baseline with brief spikes.

**Figure 2:**
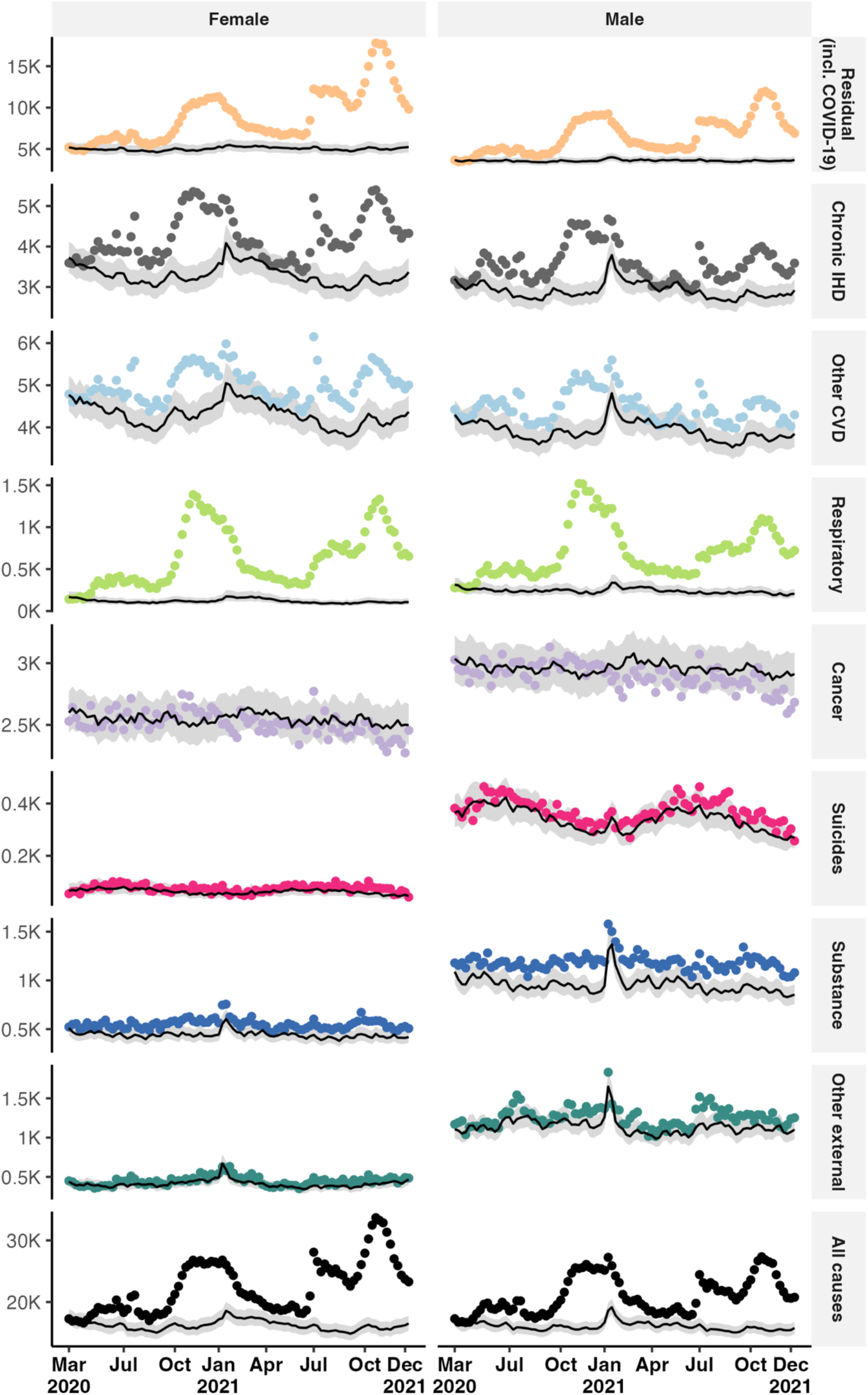
Observed (dots) and Forecasted (line) Deaths by Cause and Sex, Russia, First Week of 2019 to Last Week of 2021

By December 2021, cumulative excess deaths are dominated by COVID-19 coded deaths (Figure 3), accounting for 58% of cumulative female excess (335,303 COVID-19 coded deaths out of 580,554 total excess deaths) and 56% of cumulative male excess (260,512 COVID-19 coded deaths out of 464,360 total excess deaths). The second-largest contribution comes from cardiovascular causes. Taken together, IHD and other CVD explain roughly 60% (264,785 total deaths) of non-COVID excess in both sexes, with the cardiovascular share accumulating steadily in the end of 2020 and continuing through the end of 2021. Respiratory causes add a smaller but distinct share concentrated in the winter months. Cancer offsets a fraction of the total with cumulative deficits of 3,203 (95% CI: −4,878 to −1,590) deaths (women) and 6,149 (95% CI: −7,888 to −4,454) deaths (men). Substance-related and other external causes contribute positive but comparatively small shares, larger for men than women.

**Figure 3:**
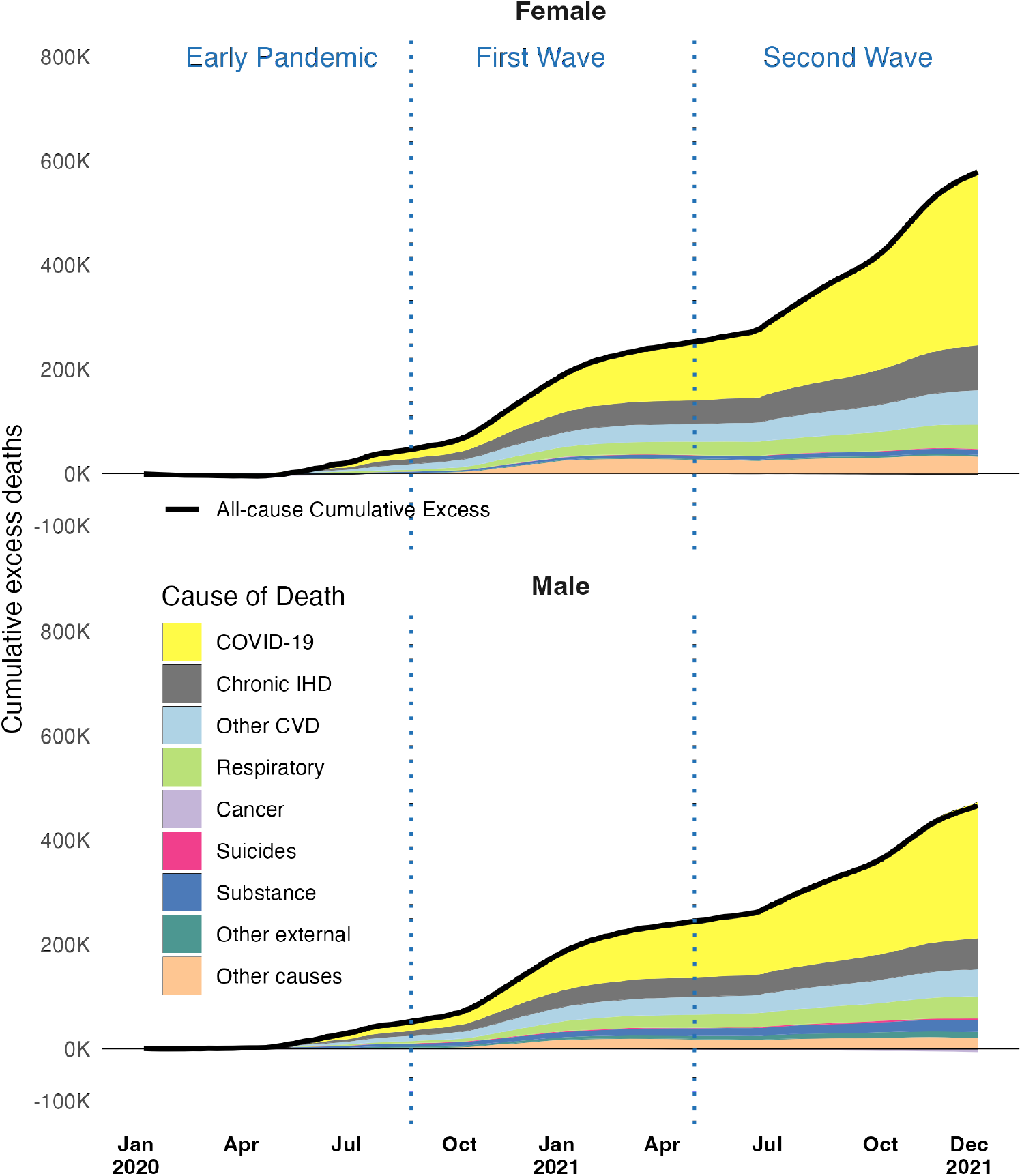
Cumulative Excess Deaths by Cause and Sex, Russia, First Week of 2020 to Last Week of 2021

Age-specific patterns sharpen the observed sex differences. In both the 45–74 and 75+ age groups, excess mortality follows a similar temporal profile — two major waves in late 2020 and late 2021 — but the magnitude is substantially larger at older ages, particularly among women (eFigure 1). Among women, cumulative excess deaths at ages 75+ (288,214) slightly exceed those at ages 45–74 (276,494), whereas among men the ordering is reversed, with larger excess at ages 45–74 than at 75+ (286,380 versus 140,978), reflecting the much smaller size of the male population at older ages.

At older ages, the COVID-19 coded component explains most of the excess for both sexes, while in 45-74 age group the non-COVID share—dominated by cardiovascular and, to a lesser extent, substance-related causes—is more prominent, particularly for men, potentially indicating a severe underreport (eFigure 2). Two features are distinctive: across the two older age groups, women exceed men in total excess, and at ages 15-44 the pattern differs, with men recording a positive excess of 37,002 deaths versus 15,856 among women.

Together, these figures indicate that (1) official COVID-19 deaths capture the timing but not the scale of the mortality shock; (2) the shortfall is primarily accounted for by cardiovascular mortality, with smaller contributions from respiratory and substance-related causes; and (3) the late 2021 wave disproportionately affected women at older ages while men accumulated larger non-COVID excess at working and older-working ages.

Turning to the regional analysis, excess mortality is highly uneven across space (Figure 4 A, Top). A broad belt of high rates runs through the central and Volga regions and into parts of the southern Urals, with several adjacent oblasts exceeding ≥ 800 per 100,000. In contrast, much of Siberia and the Far East show substantially lower rates, and several northern territories remain at the bottom of the distribution (≤ 300 per 100,000). The pattern is spatially clustered rather than random, suggesting shared exposure and health-system conditions within neighbouring regions.

**Figure 4:**
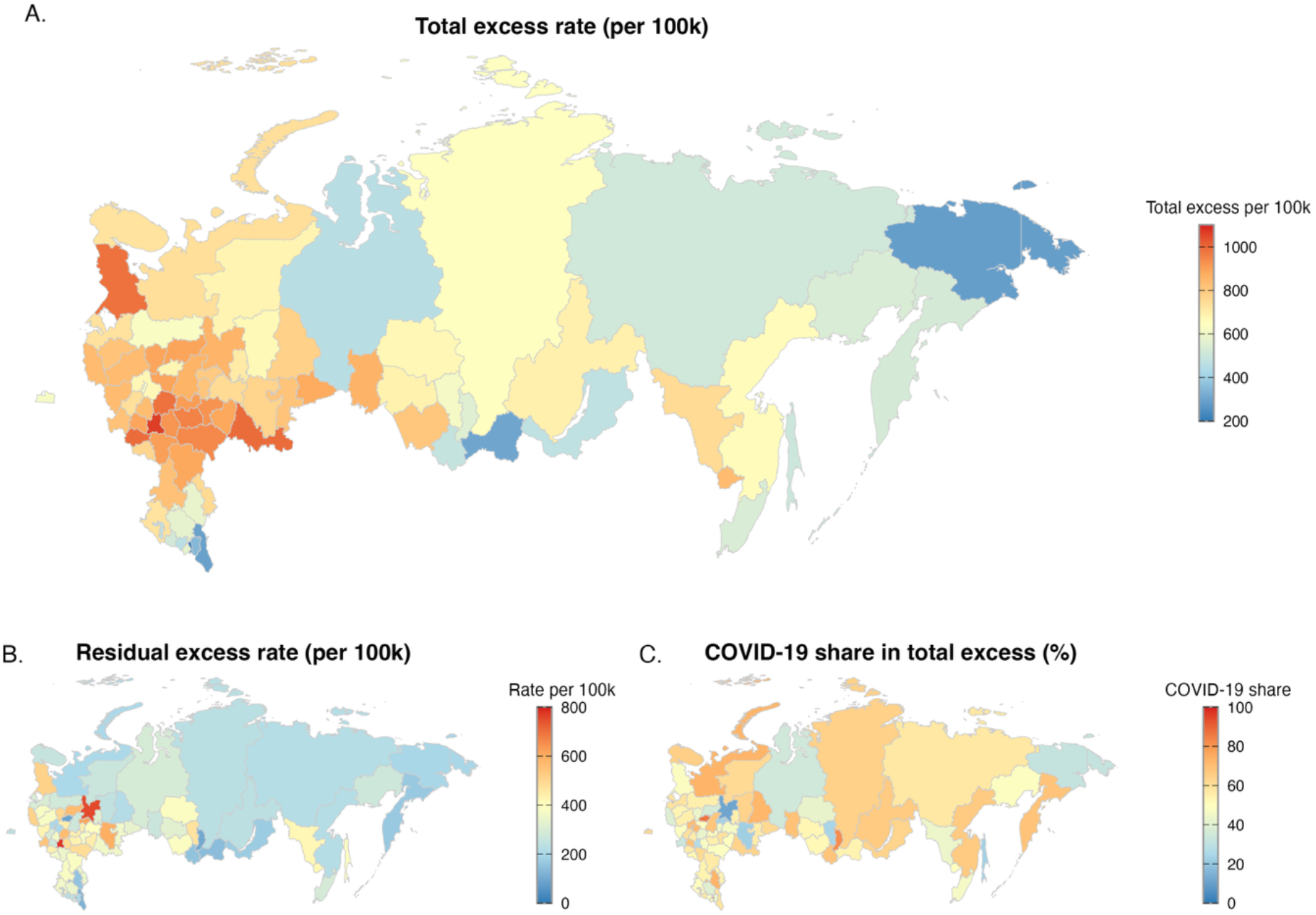
Total Excess (Top)and Residual Excess (Bottom Left) Deaths Rates, COVID-19 Share in Total Excess (Bottom Right) by Regions, Russia, Mar 2020 to Dec 2021 Note: Rates are standardised using the mean 2020-2021 population; the residual is total excess minus officially coded COVID-19 mortality and thus reflects both under-assignment and indirect deaths.

The COVID-19 map (Figure 4 C, Bottom Right) explains only part of the spatial gradient in total excess. In many of the high-excess areas, the share of officially assigned COVID-19 is elevated yet still leaves a sizeable residual component. Residual excess (Figure 4 B, Bottom Left) is concentrated in a cluster of central-southern regions, with Lipetsk standing out with an overall excess rate of 1,065.4 deaths per 100,000, its residual excess alone reaches 965.7 deaths per 100,000, and officially registered COVID-19 deaths account for only about 27% of total excess - indicating substantial under-assignment (eTable 3). Elsewhere, COVID-19 and total excess align closely and residual excess is within 0-300 per 100,000. This suggests comparatively more comprehensive assignment, but COVID-19 registrations still fall well short of total excess.

The contrast between the COVID-19 and residual panels points to heterogeneity in coding practices and/or indirect mortality burdens (e.g., cardiovascular and care-disruption effects) rather than a uniform national pattern. When mapped in counts (eFigure 4), excess deaths are dominated by a handful of populous regions. The largest metropolitan agglomerations and their surrounding oblasts account for a disproportionate share of national excess deaths—together the top five regions make up about 25% of the country’s total. Moscow alone recorded approximately 82,926 excess deaths, and Saint Petersburg a further 39,436, illustrating how population concentration heavily shapes the national burden. Conversely, many sparsely populated regions in Siberia and the Far East contribute little to the national count despite non-trivial rates. For instance, Chukotka shows an excess rate of 287.4 per 100,000 but only about 138 (statistically insignificant) excess deaths in absolute terms, reflecting its population of roughly 48,000. This divergence between rates and counts underscores two features of the Russian pandemic: concentrated demographic weight in a few regions and wide regional variation in cause assignment and mortality intensity.

## Discussion

Russia’s pandemic mortality conforms to, and intensifies, Europe’s East-West health divide. Using granular national and regional data, we document two major surges: in late 2020 and late 2021. Total excess deaths are estimated at approximately one million, yet only about half were officially assigned to COVID-19 as the underlying condition. This discrepancy is consistent with earlier evidence that COVID-19 death counts substantially understate Russia’s overall mortality burden (Kobak, 2021; Wang et al., 2022) and likely reflects the country’s comparatively narrow, autopsy-centred cause-of-death certification practices (Ivanova & Tsvetkova, 2020). At the same time, it situates Russia within a broader regional context of severe pandemic-related life-expectancy losses across Central and Eastern Europe during 2020–2021 (Aburto, Schöley, Kashnitsky, & Kashyap, 2022; Islam et al., 2021).

To understand the sources of this discrepancy, cause-of-death decomposition provides insight into the mechanisms underlying the excess mortality we estimate. Mortality from ischaemic heart disease and other cardiovascular diseases rose above pre-pandemic baselines during late 2020 and continued to accumulate through 2021, together accounting for roughly half of the non-COVID excess. Given Russia’s restrictive certification, the magnitude and timing of these increases are difficult to reconcile with indirect effects alone. A substantial share of COVID-19 deaths was likely reassigned to IHD/CVD, *alongside* genuine indirect effects (missed treatment, delayed emergency response, or reduced chronic-disease management) (Roth et al., 2022). Parallel increases in respiratory mortality further support this interpretation, as they align temporally with COVID-19 waves and are unlikely to be explained by indirect pathways alone. Taken together, the composition and timing of excess deaths indicate that certification practices materially shaped the official COVID-19 series without fully capturing the scale of the mortality shock.

These patterns must be interpreted in light of Russia’s approach to COVID-19 certification, which relied heavily on post-mortem confirmation and comparatively narrow diagnostic criteria, diverging from WHO guidance that recommends inclusion of clinically compatible and probable cases (*WHO*, 2020). Autopsy-based ascertainment is often viewed as a strength of vital statistics systems, as post-mortem examination can improve diagnostic accuracy and reduce misclassification in routine settings (Burton & Underwood, 2007). However, during a rapidly spreading respiratory epidemic, strict reliance on autopsy confirmation may introduce its own distortions. When certification is closely tied to post-mortem verification, deaths occurring outside settings where autopsies are feasible, or during periods of system strain, may be assigned to underlying chronic conditions rather than to COVID-19. In such contexts, extensive reliance on post-mortem confirmation can narrow the operational definition of pandemic mortality, potentially suppressing official COVID-19 counts while shifting deaths toward cardiovascular or other categories. In a system with constrained resources, extensive reliance on post-mortem confirmation may also have diverted personnel and facilities away from acute care, thereby compounding both misclassification and mortality.

Against this institutional backdrop, the age-sex distribution of excess mortality also reveals important patterns. Women experienced greater excess mortality at older ages (75+ and, to a comparable extent, 45–74), while men accumulated more excess deaths at younger and older-working ages. These patterns partly reflect Russia’s pre-pandemic demographic structure (notably the much smaller male population at older ages) and its long-standing male mortality disadvantage at working ages driven by cardiovascular and substance-related risks (Danilova et al., 2020; Kossova et al., 2020). Among women, the heavier toll at older ages likely reflects a combination of demographic composition, survival into older ages, and sustained exposure to health-system strain during 2021. Among men, elevated non-COVID excess at ages 15–44 and 45–74 aligns with established risk profiles and with pathways linking acute stress, substance use, and cardiovascular events.

Beyond cardiovascular mortality, patterns across other causes were more muted. Suicide mortality shows little deviation from pre-pandemic trends, echoing international evidence that suicide rates did not uniformly increase during the first years of the pandemic (Pathirathna et al., 2022; Pirkis et al., 2022; Yan et al., 2023). Cancer mortality remains flat or slightly below expected levels, a pattern that may reflect a combination of coding displacement, delayed diagnosis, and altered care-seeking behaviour rather than a true reduction in cancer incidence, and is broadly consistent with international findings (Wells & Galvani, 2022).

The structure and timing of excess mortality indicate that multiple interacting factors jointly shaped the pandemic mortality burden. The second and larger surge in late 2021 coincided with periods of extreme heat in parts of Russia^6^, a known risk factor for cardiovascular mortality that may interact with infectious disease stress and health-system strain. This coincidence suggests that environmental stressors may have intensified pandemic mortality, particularly at older ages, rather than acting independently (Grishkin, 2021).

Marked spatial variation reveals that the pandemic shock unfolded unevenly across Russia. A broad belt of high excess mortality extends through the central and Volga regions into parts of the southern Urals, while much of Siberia and the Far East exhibit lower rates. Although this configuration departs from the classic pre-pandemic southwest–northeast gradient, it nonetheless echoes long-standing regional inequalities in population health and healthcare capacity (Timonin et al., 2020, 2022). Differences between total excess mortality and officially assigned COVID-19 deaths vary widely across regions, pointing to heterogeneity in coding practices and/or indirect mortality burdens: for example, Lipetsk records very high excess yet a low COVID-19 share, implying extensive under-assignment; elsewhere, COVID-19 and total excess align more closely, though COVID-19 registrations still fall well short of all-cause excess. Such heterogeneity hints at differences in administrative capacity and coding practice differences across regions.

These outcomes unfolded within a policy and institutional context characterised by limited and short-lived non-pharmaceutical interventions, slow vaccine uptake, and persistent trust deficits - features closely associated with the East–West divide observed across Europe (Bonnet et al., 2024; Lamberova & Sonin, 2023; Wang et al., 2022). Mass vaccination scaled up gradually through 2021 but remained modest overall, reflecting scepticism, uneven delivery, and regional variation in implementation (Roshchina et al., 2022). Political and fiscal priorities further constrained sustained mitigation and likely diverted attention and capacity from pandemic control (Lamberova & Sonin, 2023; Peshkovskaya & Galkin, 2023).

Looking ahead, Russia’s mortality path will hinge on whether long-promised gains in cardiovascular care actually take hold. Since the 1970s-1990s, progress on hypertension control, secondary prevention (statins, antiplatelets), and acute cardiac care has been uneven, and alcohol policy still shapes working-age deaths (Grigoriev et al., 2014; lakunchykova et al., 2020; Nikoloski et al., 2023). COVID-19 has added backlogs in routine care, possible long-COVID effects, and extra strain on emergency services. The military conflict beginning in 2022 has introduced additional pressures on healthcare systems and mortality dynamics, further complicating recovery trajectories (Leon et al., 2022). Avoiding a lasting step-up in death rates will require steady investment in cardiovascular prevention and treatment, consistent alcohol-control measures, higher and more trusted vaccination, and basic administrative fixes to surveillance and certification. Without these, the pandemic shock risks pushing CVD mortality to a new, higher level rather than back to pre-pandemic trends. These considerations are particularly relevant in light of the federal target to raise life expectancy to 80 years by 2030 (Russian Federation. President, 2020). Recent estimates point to renewed stagnation or decline in life expectancy after the immediate post-pandemic increase; life expectancy at birth in 2024 is estimated at 72.8 years, about 0.6 years lower than in the preceding year (Scherbakova, 2025). Achieving sustained gains will therefore depend not only on recovery from the COVID-19 shock but also on structural improvements in cardiovascular prevention, healthcare delivery, and public trust.

Overall, in a population with heavy pre-existing cardiovascular risk, this combination of mistrust, hesitant vaccination, heterogeneous regional response, and restrictive certification created conditions in which the second surge was especially damaging. The resulting excess-over one million deaths in 2020-2021-constitutes a very large, and to a significant extent preventable, burden.

Our study is not without limitations. While compositional modelling helps reconcile totals and causes, it provides wide confidence intervals, that are especially noticeable on a region level. Second, our data end in 2021, limiting our capacity to evaluate subsequent changes in surveillance, vaccination, or care delivery. Additionally, our analyses use fixed-denominator regional rates (set to the mean 2020-2021 population) which improves comparability but does not capture short-term migration or mortality selection. Fourth, registration lags and reporting spikes can affect weekly patterns; we addressed these with diagnostics and sensitivity checks, but some residual bias may remain.

Despite these caveats, our approach offers notable strengths: coherent estimation of totals and causes in a single framework; weekly granularity for age- and sex-specific dynamics; and a transparent separation of officially coded COVID-19 from the residual. Substantively, the findings point to several implications. First, where certification is restrictive, cause-specific excess—particularly cardiovascular—should be tracked alongside official COVID-19 counts as divergences between the two may signal misclassification or indirect effects rather than statistical noise. Second, continuity of cardiovascular care during respiratory epidemics is critical: maintaining emergency response, medications, and outpatient management may mitigate large non-COVID excess. Third, regional targeting matters. Clusters with high residual excess are candidates for *audit of certification practices* and possibly for investment in health-system capacity.

In sum, Russia’s pandemic mortality was not only large but also compositionally skewed toward cardiovascular deaths, with sex, age, and regional patterns that challenge simple readings of official COVID-19 statistics. Excess-mortality frameworks that integrate totals with cause composition-such as the one used here - are well suited to countries with heterogeneous coding practices and large chronic-disease burdens, and they highlight the value of timely, weekly cause-of-death data for public health surveillance and accountability.

## Supporting information

Online-Only Figures and Tables

## Data Availability

Materials for reproduction, including scripts, will be available at peer review and upon publication.

## Acknowledgements

Previous version of this work was presented at the Population Association America conference in 2025, Washington DC. This work has benefited from the helpful feedback of the LCDS population health working group. ED and AT were supported by the Leverhulme Trust (Grant RC-2018-003) for the Leverhulme Centre for Demographic Science. Sergey ST was supported by the MRFF Cardiovascular Mission Grant (2034149). JMA was supported by a Wellcome Trust Career Development Award (07859/Z/23/Z). ED was supported by the EPSRC Centre for Doctoral Training in Healthcare Data Science (EP/Y035321/1). The views expressed in this article are those of the authors and do not necessarily represent the views of any funding agency.

## Footnotes

1 In 2021, the age-standardised CVD mortality accounted for 395 per 100,000 (GBD, 2021)

2 Russian Federation. (2021). World Health Organization. https://data.who.int/countries/643

3 ICD-10 codes for causes are available in Supplementary material eTable1

4 See Supplementary material eTable 2 for performance metrics

5 See eFigure 3 in the Supplementary Material for the same graph extended to the full pre-pandemic (training) period to assess model fit.

6 Heat wave in Russia brings record-breaking temperatures north of Arctic Circle - ABC News. June 2021. Accessed November 28, 2025. https://abcnews.go.com/International/heat-wave-russia-brings-record-breaking-temperatures-north/story?id=78446355#

## References

Aburto, J. M., Kashyap, R., Schöley, J., Angus, C., Ermisch, J., Mills, M. C., & Dowd, J. B. (2021). Estimating the burden of the COVID-19 pandemic on mortality, life expectancy and lifespan inequality in England and Wales: A population-level analysis. Journal of Epidemiology and Community Health, 75(8), 735–740. 10.1136/jech-2020-215505

Aburto, J. M., Schöley, J., Kashnitsky, I., & Kashyap, R. (2022). Life expectancy declines in Russia during the COVID-19 pandemic in 2020. International Journal of Epidemiology, 51(5), 1695–1697. 10.1093/ije/dyac055

Aburto, J. M., Schöley, J., Kashnitsky, I., Zhang, L., Rahal, C., Missov, T. I., Mills, M. C., Dowd, J. B., & Kashyap, R. (2022a). Quantifying impacts of the COVID-19 pandemic through life-expectancy losses: A population-level study of 29 countries. International Journal of Epidemiology, 51(1), 63–74. 10.1093/ije/dyab207

Aburto, J. M., Schöley, J., Kashnitsky, I., Zhang, L., Rahal, C., Missov, T. I., Mills, M. C., Dowd, J. B., & Kashyap, R. (2022b). Quantifying impacts of the COVID-19 pandemic through life-expectancy losses: A population-level study of 29 countries. International Journal of Epidemiology, 51(1), 63–74. 10.1093/ije/dyab207

Aleksandrova, G. A., Golubev, N. A., Tiurina, E. M., Ogryzko, E. V., & Shelepova, E. A. (2019). Resources and activities of medical organizations in the healthcare system: Medical personnel. Part 1. Report of the Ministry of Health. https://minzdrav.gov.ru/ministry/61/22/stranitsa-979/statisticheskie-i-informatsionnye-materialy/statisticheskiy-sbornik-2018-god(open in a new window).

Alizadeh, H., Sharifi, A., Damanbagh, S., Nazarnia, H., & Nazarnia, M. (2023). Impacts of the COVID-19 pandemic on the social sphere and lessons for crisis management: A literature review. Natural Hazards, 1–26. 10.1007/s11069-023-05959-2

Avena, N. M., Simkus, J., Lewandowski, A., Gold, M. S., & Potenza, M. N. (2021). Substance Use Disorders and Behavioral Addictions During the COVID-19 Pandemic and COVID-19-Related Restrictions. Frontiers in Psychiatry, 12, 653674. 10.3389/fpsyt.2021.653674

Beaney, T., Clarke, J. M., Jain, V., Golestaneh, A. K., Lyons, G., Salman, D., & Majeed, A. (2020). Excess mortality: The gold standard in measuring the impact of COVID-19 worldwide? Journal of the Royal Society of Medicine, 113(9), 329–334. 10.1177/0141076820956802

Bergeron-Boucher, M.-P., Canudas-Romo, V., Oeppen, J. E., & Vaupel, J. W. (2017). Coherent forecasts of mortality with compositional data analysis. Demographic Research, 37, 527–566. 10.4054/DemRes.2017.37.17

Blackburn, M., Hutcheson, D., Tsumarova, E., & Petersson, B. (2023). Covid-19 and the Russian Regional Response: Blame Diffusion and Attitudes to Pandemic Governance. Canadian Journal of European and Russian Studies, 16(1), 29–54. 10.22215/cjers.v16i1.3955

Bonnet, F., Grigoriev, P., Sauerberg, M., Alliger, I., Mühlichen, M., & Camarda, C.-G. (2024). Spatial disparities in the mortality burden of the covid-19 pandemic across 569 European regions (2020-2021). Nature Communications, 15(1), 4246. 10.1038/s41467-024-48689-0

Buera, F., Fattal-Jaef, R., Hopenhayn, H., Neumeyer, P. A., & Shin, Y. (2021). The Economic Ripple Effects of COVID-19 (W28704; p. w28704). National Bureau of Economic Research. 10.3386/w28704

Burki, T. K. (2020). The Russian vaccine for COVID-19. The Lancet Respiratory Medicine, 8(11), e85–e86. 10.1016/S2213-2600(20)30402-1

Burton, J. L., & Underwood, J. (2007). Clinical, educational, and epidemiological value of autopsy. The Lancet, 369(9571), 1471–1480. 10.1016/S0140-6736(07)60376-6

Chen, R., Charpignon, M.-L., Raquib, R. V., Wang, J., Meza, E., Aschmann, H. E., DeVost, M. A., Mooney, A., Bibbins-Domingo, K., Riley, A. R., Kiang, M. V., Chen, Y.-H., Stokes, A. C., & Glymour, M. M. (2023). Excess Mortality With Alzheimer Disease and Related Dementias as an Underlying or Contributing Cause During the COVID-19 Pandemic in the US. JAMA Neurology. 10.1001/jamaneurol.2023.2226

Chetty, R., Friedman, J. N., Stepner, M., & Opportunity Insights Team. (2024). The Economic Impacts of COVID-19: Evidence from a New Public Database Built Using Private Sector Data*. The Quarterly Journal of Economics, 139(2), 829–889. 10.1093/qje/qjad048

Clair, R., Gordon, M., Kroon, M., & Reilly, C. (2021). The effects of social isolation on well-being and life satisfaction during pandemic. Humanities and Social Sciences Communications, 8(1), Article 1. 10.1057/s41599-021-00710-3

Danilova, I., Shkolnikov, V. M., Andreev, E., & Leon, D. A. (2020). The changing relation between alcohol and life expectancy in Russia in 1965–2017. Drug and Alcohol Review, 39(7), 790–796. 10.1111/dar.13034

Degtiareva, E., Tilstra, A. M., Schöley, J., Kashyap, R., & Dowd, J. B. (2024). Cause-Specific Excess Mortality in the US During the COVID-19 Pandemic. 10.1101/2024.03.05.24303783

Demertzis, N., & Klironomos, N. (2026). Trusting during the pandemic: The effect of political, social and scientific trust on COVID-19 mortality across Europe. Frontiers in Political Science, 7. 10.3389/fpos.2025.1678788

Dyer, O. (2020). Covid-19: Russia admits to understating deaths by more than two thirds. BMJ, m4975. 10.1136/bmj.m4975

Dyer, O. (2021). Covid-19: Russia’s statistics agency reports much higher death toll than country’s health ministry. BMJ, 372, n440. 10.1136/bmj.n440

Fadlallah, R., El-Jardali, F., Kalach, N., Karroum, L. B., Hoteit, R., Aoun, A., Al-Hakim, L., Verdugo-Paiva, F., Rada, G., Fretheim, A., Lewin, S., Ludolph, R., & Akl, E. A. (2023). Public Health and Social Measures (PHSM) interventions to control COVID-19 An Overview of Systematic Reviews (p. 2023.11.21.23298387). medRxiv. 10.1101/2023.11.21.23298387

Fekadu, G., Bekele, F., Tolossa, T., Fetensa, G., Turi, E., Getachew, M., Abdisa, E., Assefa, L., Afeta, M., Demisew, W., Dugassa, D., Diriba, D. C., & Labata, B. G. (2021). Impact of COVID-19 pandemic on chronic diseases care follow-up and current perspectives in low resource settings: A narrative review. International Journal of Physiology, Pathophysiology and Pharmacology, 13(3), 86–93.

Grigoriev, P., & Andreev, E. M. (2015). The Huge Reduction in Adult Male Mortality in Belarus and Russia: Is It Attributable to Anti-Alcohol Measures? PLOS ONE, 10(9), e0138021. 10.1371/journal.pone.0138021

Grigoriev, P., Meslé, F., Shkolnikov, V. M., Andreev, E., Fihel, A., Pechholdova, M., & Vallin, J. (2014). The Recent Mortality Decline in Russia: Beginning of the Cardiovascular Revolution? Population and Development Review, 40(1), 107–129. 10.1111/j.1728-4457.2014.00652.x

Grishkin, D. (2021, July 13). Russia Marks Second-Hottest June in History With More Record Heat to Come – Weather Chief. The Moscow Times. https://www.themoscowtimes.com/2021/07/13/russia-marks-second-hottest-june-in-history-with-more-record-heat-to-come-weather-chief-a74510

Hajdu, T., Krekó, J., & Tóth, C. G. (2024). Inequalities in regional excess mortality and life expectancy during the COVID-19 pandemic in Europe. Scientific Reports, 14(1), 3835. 10.1038/s41598-024-54366-5

Hastie, T., & Tibshirani, R. (1986). Generalized Additive Models. Statistical Science, 1(3). 10.1214/ss/1177013604

International Guidelines for Certification and Classification (Coding) of COVID-19 as Cause of Death. (2020). World Health Organization. https://www.who.int/publications/m/item/international-guidelines-for-certification-and-classification-(coding)-of-covid-19-as-cause-of-death

Islam, N., Jdanov, D. A., Shkolnikov, V. M., Khunti, K., Kawachi, I., White, M., Lewington, S., & Lacey, B. (2021). Effects of covid-19 pandemic on life expectancy and premature mortality in 2020: Time series analysis in 37 countries. BMJ, e066768. 10.1136/bmj-2021-066768

Ivanova, P., & Tsvetkova, M. (2020, May 19). Russia says many coronavirus patients died of other causes. Some disagree. Reuters. https://www.reuters.com/article/us-health-coronavirus-russia-casualties-idUSKBN22V1Q7

Karlinsky, A., & Kobak, D. (2021). The World Mortality Dataset: Tracking excess mortality across countries during the COVID-19 pandemic [Preprint]. Epidemiology. 10.1101/2021.01.27.21250604

King, E. J., & Dudina, V. I. (2021). COVID-19 in Russia: Should we expect a novel response to the novel coronavirus? Global Public Health, 16(8–9), 1237–1250. 10.1080/17441692.2021.1900317

Kjærgaard, S., Ergemen, Y. E., Kallestrup-Lamb, M., Oeppen, J., & Lindahl-Jacobsen, R. (2019). Forecasting Causes of Death by Using Compositional Data Analysis: The Case of Cancer Deaths. Journal of the Royal Statistical Society Series C: Applied Statistics, 68(5), 1351–1370. 10.1111/rssc.12357

Kobak, D. (2021). Excess Mortality Reveals Covid’s True Toll in Russia. Significance, 18(1), 16–19. 10.1111/1740-9713.01486

Kontoangelos, K., Economou, M., & Papageorgiou, C. (2020). Mental Health Effects of COVID-19 Pandemia: A Review of Clinical and Psychological Traits. Psychiatry Investigation, 17(6), 491–505. 10.30773/pi.2020.0161

lakunchykova, O., Averina, M., Wilsgaard, T., Watkins, H., Malyutina, S., Ragino, Y., Keogh, R. H., Kudryavtsev, A. V., Govorun, V., Cook, S., Schirmer, H., Eggen, A. E., Hopstock, L. A., & Leon, D. A. (2020). Why does Russia have such high cardiovascular mortality rates? Comparisons of blood-based biomarkers with Norway implicate non-ischaemic cardiac damage. Journal of Epidemiology and Community Health, 74(9), 698–704. 10.1136/jech-2020-213885

Lamberova, N., & Sonin, K. (2023). Information Manipulation and Repression: A Theory and Evidence from the COVID Response in Russia (SSRN Scholarly Paper 4174501). 10.2139/ssrn.4174501

Lazarus, J. V., Ratzan, S. C., Palayew, A., Gostin, L. O., Larson, H. J., Rabin, K., Kimball, S., & El-Mohandes, A. (2021). A global survey of potential acceptance of a COVID-19 vaccine. Nature Medicine, 27(2), 225–228. 10.1038/s41591-020-1124-9

Leon, D. A., Jdanov, D. A., & Shkolnikov, V. M. (2019). Trends in life expectancy and age-specific mortality in England and Wales, 1970–2016, in comparison with a set of 22 high-income countries: An analysis of vital statistics data. The Lancet Public Health, 4(11), e575–e582. 10.1016/S2468-2667(19)30177-X

Leon, D. A., Jdanov, D., Gerry, C. J., Grigoriev, P., Jasilionis, D., McKee, M., Meslé, F., Penina, O., Twigg, J., Vallin, J., & Vågerö, D. (2022). The Russian invasion of Ukraine and its public health consequences. The Lancet Regional Health - Europe, 15, 100358. 10.1016/j.lanepe.2022.100358

Levitt, M., Zonta, F., & Ioannidis, J. P. A. (2022). Comparison of pandemic excess mortality in 2020–2021 across different empirical calculations. Environmental Research, 213, 113754. 10.1016/j.envres.2022.113754

Lopez-Leon, S., Wegman-Ostrosky, T., Perelman, C., Sepulveda, R., Rebolledo, P. A., Cuapio, A., & Villapol, S. (2021). More than 50 long-term effects of COVID-19: A systematic review and meta-analysis. Scientific Reports, 11(1), 16144. 10.1038/s41598-021-95565-8

Makarova, M. N., Pyshmintseva, O. A., & Institute of Economics, Ural Branch of the Russian Academy of Sciences, Ekaterinburg, Russian Federation. (2021). Excess mortality in Russian regions during the COVID-19 pandemic. R-Economy, 7(4), 225–234. 10.15826/recon.2021.7.4.020

Matskeplishvili, S., & Kontsevaya, A. (2021). Cardiovascular Health, Disease, and Care in Russia. Circulation, 144(8), 586–588. 10.1161/CIRCULATIONAHA.121.055239

Moynihan, R., Sanders, S., Michaleff, Z. A., Scott, A. M., Clark, J., To, E. J., Jones, M., Kitchener, E., Fox, M., Johansson, M., Lang, E., Duggan, A., Scott, I., & Albarqouni, L. (2021). Impact of COVID-19 pandemic on utilisation of healthcare services: A systematic review. BMJ Open, 11(3), e045343. 10.1136/bmjopen-2020-045343

Msemburi, W., Karlinsky, A., Knutson, V., Aleshin-Guendel, S., Chatterji, S., & Wakefield, J. (2023). The WHO estimates of excess mortality associated with the COVID-19 pandemic. Nature, 613(7942), 130–137. 10.1038/s41586-022-05522-2

Nicola, M., Alsafi, Z., Sohrabi, C., Kerwan, A., Al-Jabir, A., Iosifidis, C., Agha, M., & Agha, R. (2020). The socio-economic implications of the coronavirus pandemic (COVID-19): A review. International Journal of Surgery, 78, 185–193. 10.1016/j.ijsu.2020.04.018

Nikoloski, Z., Shkolnikov, V. M., & Mossialos, E. (2023). Preventable mortality in the Russian Federation: A retrospective, regional level study. The Lancet Regional Health - Europe, 29, 100631. 10.1016/j.lanepe.2023.100631

Oeppen, J. (2008). Coherent forecasting of multiple-decrement life tables: A test using Japanese cause of death data. European Population Conference.

Pathirathna, M. L., Nandasena, H. M. R. K. G., Atapattu, A. M. M. P., & Weerasekara, I. (2022). Impact of the COVID-19 pandemic on suicidal attempts and death rates: A systematic review. BMC Psychiatry, 22(1), 506. 10.1186/s12888-022-04158-w

Peshkovskaya, A., & Galkin, S. (2023). Health behavior in Russia during the COVID-19 pandemic. Frontiers in Public Health, 11, 1276291. 10.3389/fpubh.2023.1276291

Pirkis, J., Gunnell, D., Shin, S., Del Pozo-Banos, M., Arya, V., Aguilar, P. A., Appleby, L., Arafat, S. M. Y., Arensman, E., Ayuso-Mateos, J. L., Balhara, Y. P. S., Bantjes, J., Baran, A., Behera, C., Bertolote, J., Borges, G., Bray, M., Brečić, P., Caine, E., … Spittal, M. J. (2022). Suicide numbers during the first 9-15 months of the COVID-19 pandemic compared with pre-existing trends: An interrupted time series analysis in 33 countries. eClinicalMedicine, 51. 10.1016/j.eclinm.2022.101573

Policy Responses to COVID-19 in the Russian Federation (World Bank Group.). (2020). http://documents.worldbank.org/curated/en/292961593661069327

Polizzi, A., Zhang, L., Timonin, S., Gupta, A., Dowd, J. B., Leon, D. A., & Aburto, J. M. (2025). Indirect effects of the COVID-19 pandemic: A cause-of-death analysis of life expectancy changes in 24 countries, 2015 to 2022. PNAS Nexus, 4(1), pgae508. 10.1093/pnasnexus/pgae508

Roshchina, Y., Roshchin, S., & Rozhkova, K. (2022). Determinants of COVID-19 vaccine hesitancy and resistance in Russia. Vaccine, 40(39), 5739–5747. 10.1016/j.vaccine.2022.08.042

Roth, G. A., Vaduganathan, M., & Mensah, G. A. (2022). Impact of the COVID-19 Pandemic on Cardiovascular Health in 2020: JACC State-of-the-Art Review. Journal of the American College of Cardiology, 80(6), 631–640. 10.1016/j.jacc.2022.06.008

Russian Federation. President. (2020, July). Decree on the national development goals of the Russian Federation through 2030 [Ukaz o natsional’nykh tseliakh razvitiia Rossiiskoi Federatsii na period do 2030 goda]. http://kremlin.ru/acts/news/63728

Scherbakova, E. (2025). The first demographic results of 2024 in Russia [Pervye Demographicheskie Itogi 2024 Goda v Rossii]. Demoscope Weekly, (1069).

Scherbov, S., Gietel-Basten, S., Ediev, D., Shulgin, S., & Sanderson, W. (2022). COVID-19 and excess mortality in Russia: Regional estimates of life expectancy losses in 2020 and excess deaths in 2021. PLOS ONE, 17(11), e0275967. 10.1371/journal.pone.0275967

Schöley, J. (2024). XCOD: Estimate expected and excess deaths by cause using coherent compositional regression. [Computer software]. https://zenodo.org/doi/10.5281/zenodo.13353995

Schöley, J., Aburto, J. M., Kashnitsky, I., Kniffka, M. S., Zhang, L., Jaadla, H., Dowd, J. B., & Kashyap, R. (2022). Life expectancy changes since COVID-19. Nature Human Behaviour, 6(12), 1649–1659. 10.1038/s41562-022-01450-3

Shchur, A., & Timonin, S. (2021). Center-peripheral differences in life expectancy in Russia: A regional analysis. Demographic Review [Demographicheskoye Obozrenie], 7(5), 63–83. 10.17323/demreview.v7i5.13198

Shkolnikov, V., Andreev, E. M., McKee, M., & Leon, D. A. (2013). Components and possible determinants of decrease in Russian mortality in 2004-2010. Demographic Research, 28, 917–950. 10.4054/DemRes.2013.28.32

Shkolnikov, V. M., Timonin, S., Jdanov, D., Islam, N., & Leon, D. A. (2023). East-West mortality disparities during the COVID-19 pandemic widen the historical longevity divide in Europe (p. 2023.11.08.23298275). medRxiv. 10.1101/2023.11.08.23298275

Sobczak, M., & Pawliczak, R. (2022). COVID-19 mortality rate determinants in selected Eastern European countries. BMC Public Health, 22(1), 2088. 10.1186/s12889-022-14567-x

Stronski, P. (2021, October). Russia’s Response to Its Spiraling COVID-19 Crisis Is Too Little, Too Late. Carnegie Endowment for International Peace. https://carnegieendowment.org/2021/10/28/russia-s-response-to-its-spiraling-covid-19-crisis-is-too-little-too-late-pub-85677

The age-standardized number of deaths from cardiovascular disease per 100,000 population in 2021. (2021). [Dataset]. GBD Compare Data Visualization. https://world-heart-federation.org/world-heart-observatory/countries/russia/

Timonin, S., Jasilionis, D., Shkolnikov, V. M., & Andreev, E. (2020). New perspective on geographical mortality divide in Russia: A district-level cross-sectional analysis, 2008–2012. Journal of Epidemiology and Community Health, 74(2), 144–150. 10.1136/jech-2019-213239

Timonin, S., Klimkin, I., Shkolnikov, V. M., Andreev, E., McKee, M., & Leon, D. A. (2022). Excess mortality in Russia and its regions compared to high income countries: An analysis of monthly series of 2020. SSM - Population Health, 17, 101006. 10.1016/j.ssmph.2021.101006

Vishnevsky, A., Andreev, E., & Timonin, S. (2017). Mortality from cardiovascular diseases and life expectancy in Russia. Демографическое Обозрение, 45–70. 10.17323/demreview.v4i5.8566

Wadhera, R. K., Shen, C., Gondi, S., Chen, S., Kazi, D. S., & Yeh, R. W. (2021). Cardiovascular Deaths During the COVID-19 Pandemic in the United States. Journal of the American College of Cardiology, 77(2), 159–169. 10.1016/j.jacc.2020.10.055

Wang, H., Paulson, K. R., Pease, S. A., Watson, S., Comfort, H., Zheng, P., Aravkin, A. Y., Bisignano, C., Barber, R. M., Alam, T., Fuller, J. E., May, E. A., Jones, D. P., Frisch, M. E., Abbafati, C., Adolph, C., Allorant, A., Amlag, J. O., Bang-Jensen, B., … Murray, C. J. L. (2022). Estimating excess mortality due to the COVID-19 pandemic: A systematic analysis of COVID-19-related mortality, 2020–21. The Lancet, 399(10334), 1513–1536. 10.1016/S0140-6736(21)02796-3

Wells, C. R., & Galvani, A. P. (2022). Impact of the COVID-19 pandemic on cancer incidence and mortality. The Lancet Public Health, 7(6), e490–e491. 10.1016/S2468-2667(22)00111-6

Yaddanapudi, L., Hahn, J., & Ladikas, M. (2023). Decreasing trust in health institutions in EU during COVID-19: A Spatio-temporal analysis. The European Journal of Public Health, 33(Suppl 2), ckad160.594. 10.1093/eurpub/ckad160.594

Yan, Y., Hou, J., Li, Q., & Yu, N. X. (2023). Suicide before and during the COVID-19 Pandemic: A Systematic Review with Meta-Analysis. International Journal of Environmental Research and Public Health, 20(4), 3346. 10.3390/ijerph20043346

Zamyatnina, E. S., Shkolnikov, V. M., & Shchur, A. E. (2025). Associations Between the Mortality Trends by Cause of Death and Hazardous Alcohol Consumption in Russia. The “Conveyor Belt Effect” in Action? Population and Economics, 9(1), 214–243. 10.3897/popecon.9.e127917

