## Supplementary material for "Beyond COVID-19 Deaths: Cause-Specific Analysis of Excess Mortality in Russia": Online-Only Figures and Tables

### Supplementary materials

#### Code availability:

All analyses were conducted in R version 4.3.0. Materials for reproduction, including metadata and scripts, will be available at peer review and upon publication.

**eTable 1:** Cause-of-Death Classification and Corresponding ICD-10 Codes

| Cause of death | ICD10 codes |
|---|---|
| Cancer | C00-D48 |
| Respiratory | J00-J22 |
| Chronic IHD | I25 |
| Rest of CVD | all of I00-I99 excluding I25 |
| Substance-related | F10-F19, I42.6, K70, K73-74, X40-45, X85, Y10-Y15 |
| Suicides | X60-X84, Y20, Y87.0 |
| Rest of external causes of death | V01-Y89, excluding those already assigned to substance-related and suicides |
| COVID-19 | U07.1, U07.2 |
| Residual | All codes not included in any category above |

**eTable 2:** Model Performance Grid

Panel A: Excess Mortality Modelling by Age, Sex and Cause of Death

| Measure/Model | Weekly Average | Serfling Model | Negative Binomial | GAM |
|---|---|---|---|---|
| MAE | 186 | 133.2 | 109.9 | 99.7 |
| RMSE | 267.8 | 218.7 | 181.4 | 156.7 |

Panel B: Excess Mortality Modelling by Region

| Measure/Model | Monthly Average | Serfling Model | Negative Binomial | GAM |
|---|---|---|---|---|
| MAE | 134.7 | 166.2 | 175.4 | 128.7 |
| RMSE | 665.1 | 850.5 | 935.1 | 659.6 |

Note: Model selection procedures, including R code and data used in the model-choice exercise will be available at peer review and upon publication.

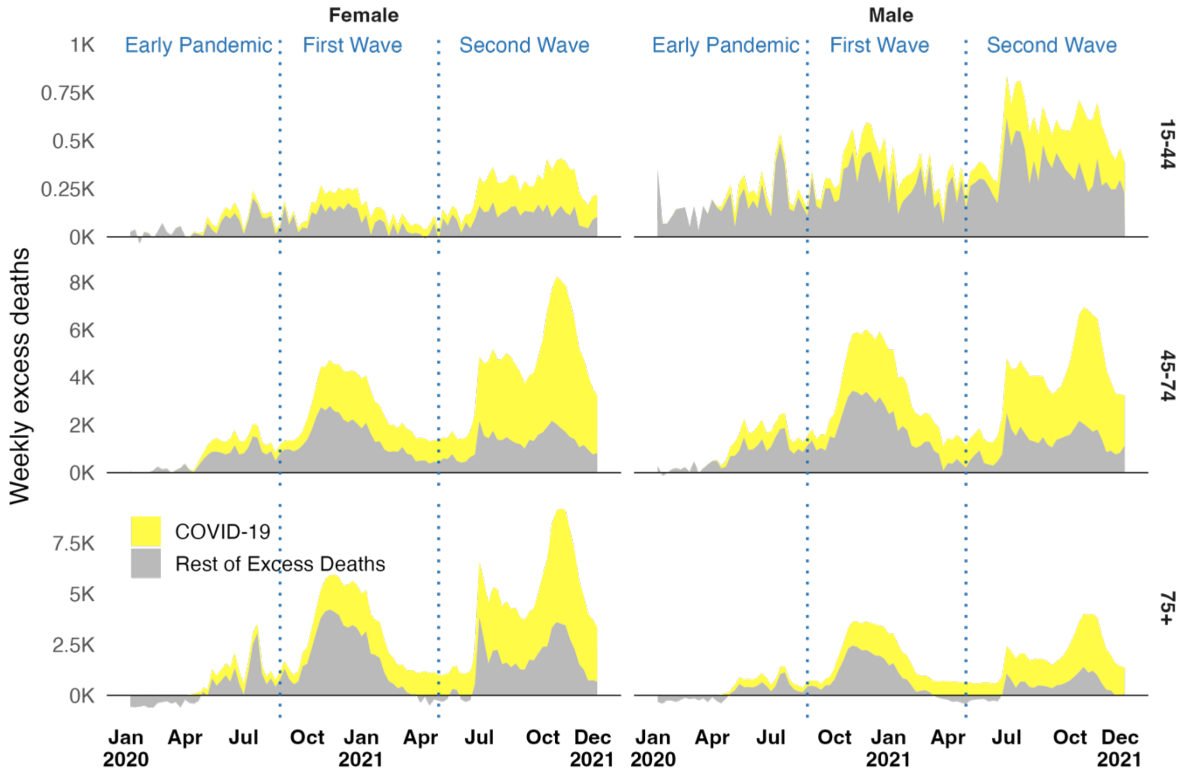


*eFigure 1: Weekly Excess Mortality by Age and Sex, Russia, First Week of 2020 to Last Week of 2021.*

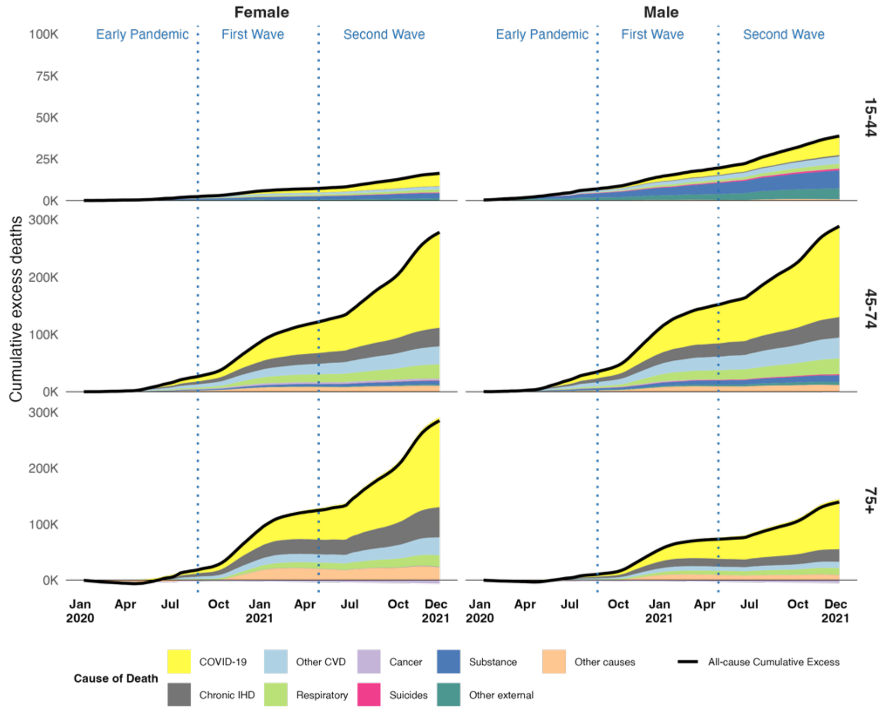


*eFigure 2: Cumulative Excess Deaths by Cause, Age and Sex, Russia, First Week of 2020 to Last Week of 2021*

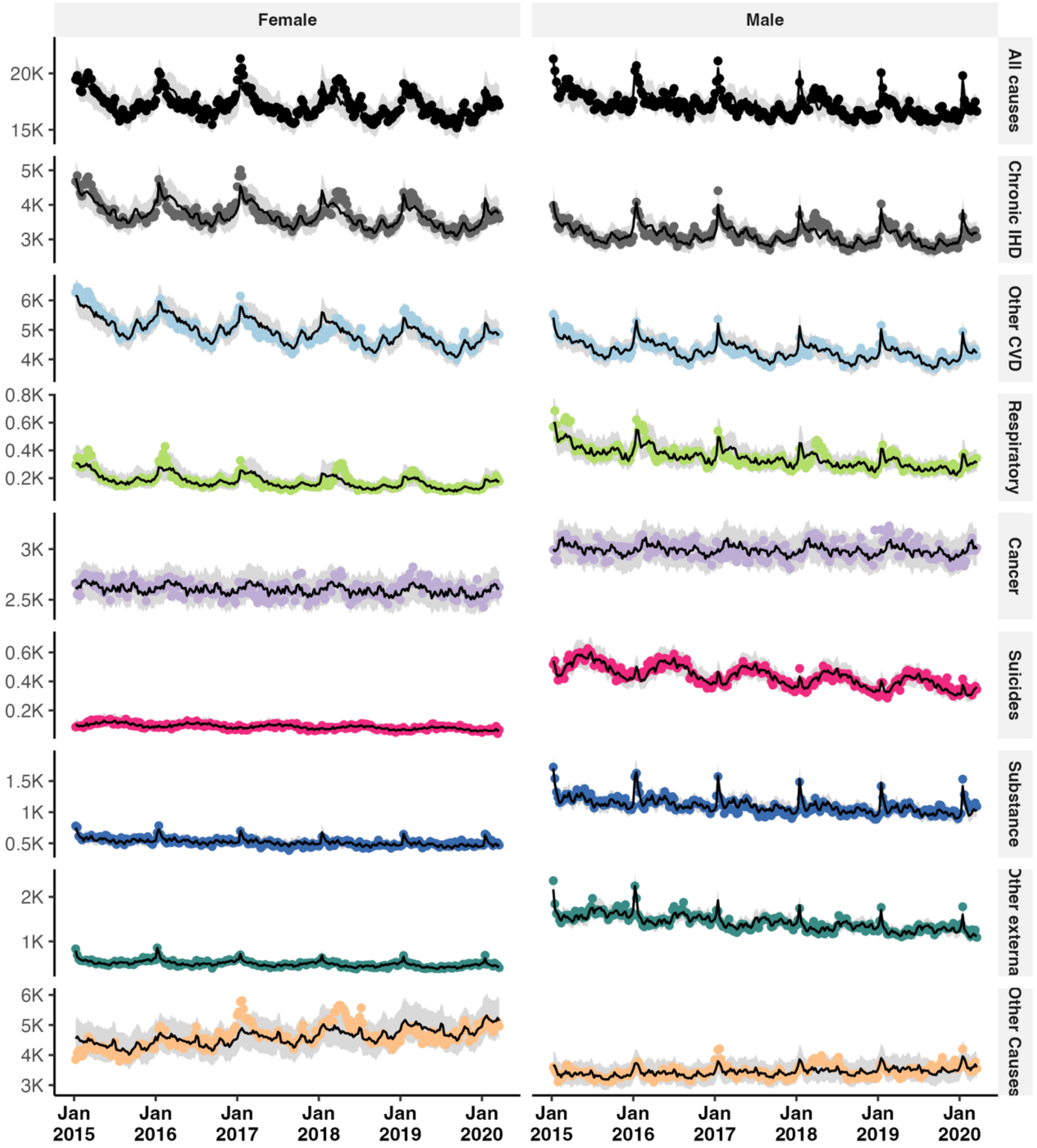


*eFigure 3: Observed (dots) and Forecasted (line) Deaths by Cause and Sex, Russia, First Week of 2014 to Last Week before Pandemic (11th Week of 2020)*

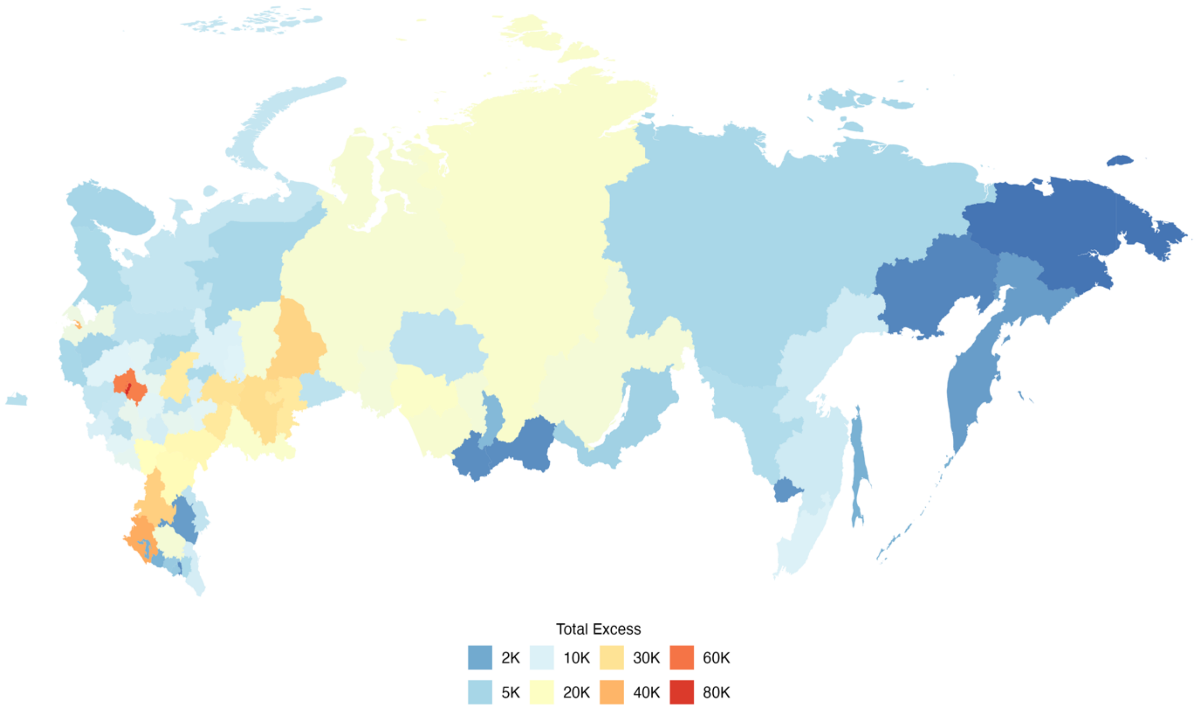


*eFigure 4: Total Absolute Excess Deaths by Region, Russia, Jan 202 to Dec 2021*

**eTable 3:** Total Excess Deaths Rates and COVID-19 Rates by Region, Russia, Mar 2020-Dec 2021

| Region | Absolute Excess | Official COVID-19 Deaths | Population | Excess Rate | COVID-19 Share in Excess |
|---|---|---|---|---|---|
| Russia | 1,047,715 | 601,668 | 144,995,964 | 722.6 | 57.4 |
| Moscow | 82,926 | 66,308 | 12,980,153 | 638.9 | 80.0 |
| Moscow.oblast | 56,833 | 40,104 | 8,447,220 | 672.8 | 70.6 |
| Krasnodar | 42,323 | 21,063 | 5,814,365 | 727.9 | 49.8 |
| Saint.Petersburg | 39,436 | 35,046 | 5,591,432 | 705.3 | 88.9 |
| Rostov | 34,674 | 18,635 | 4,213,749 | 822.9 | 53.7 |
| Sverdlovsk | 33,393 | 24,127 | 4,288,198 | 778.7 | 72.2 |
| Bashkortostan | 31,483 | 7,073 | 4,099,168 | 768.0 | 22.5 |
| Tatarstan | 30,009 | 14,007 | 3,997,965 | 750.6 | 46.7 |
| Samara | 28,246 | 14,181 | 3,184,593 | 887.0 | 50.2 |
| Chelyabinsk | 28,025 | 15,622 | 3,445,338 | 813.4 | 55.7 |

| Region | Absolute Excess | Official COVID-19 Deaths | Population | Excess Rate | COVID-19 Share in Excess |
|---|---|---|---|---|---|
| Nizhny.Novgorod | 27,003 | 18,344 | 3,143,028 | 859.1 | 67.9 |
| Saratov | 23,188 | 11,282 | 2,456,095 | 944.1 | 48.6 |
| Volgograd | 22,201 | 12,462 | 2,511,585 | 884.0 | 56.1 |
| Voronezh | 20,924 | 12,878 | 2,319,317 | 902.1 | 61.5 |
| Novosibirsk | 19,246 | 10,187 | 2,803,135 | 686.6 | 52.9 |
| Orenburg | 18,688 | 11,877 | 1,878,217 | 995.0 | 63.5 |
| Krasnoyarsk | 18,420 | 11,826 | 2,863,308 | 643.3 | 64.2 |
| Altai.Krai | 17,686 | 9,205 | 2,191,276 | 807.1 | 52.0 |
| Tyumen | 17,277 | 6,041 | 3,798,430 | 454.8 | 35.0 |
| Perm | 17,026 | 10,820 | 2,550,342 | 667.6 | 63.5 |
| Stavropol | 16,732 | 6,692 | 2,903,741 | 576.2 | 40.0 |
| Irkutsk | 16,439 | 10,854 | 2,380,340 | 690.6 | 66.0 |
| Kemerovo | 16,198 | 3,920 | 2,621,070 | 618.0 | 24.2 |
| Omsk | 16,116 | 11,110 | 1,877,853 | 858.2 | 68.9 |
| Leningrad | 14,452 | 8,494 | 1,977,890 | 730.7 | 58.8 |
| Tula | 12,595 | 7,055 | 1,507,735 | 835.4 | 56.0 |
| Lipetsk | 12,249 | 3,323 | 1,149,766 | 1065.4 | 27.1 |
| Penza | 12,225 | 6,707 | 1,278,313 | 956.4 | 54.9 |
| Vladimir | 12,151 | 6,134 | 1,358,521 | 894.4 | 50.5 |
| Belgorod | 11,799 | 6,203 | 1,545,080 | 763.6 | 52.6 |
| Ulyanovsk | 11,159 | 6,577 | 1,206,390 | 925.0 | 58.9 |
| Ryazan | 10,924 | 4,667 | 1,109,828 | 984.3 | 42.7 |
| Yaroslavl | 10,875 | 4,731 | 1,221,156 | 890.5 | 43.5 |
| Kursk | 10,854 | 6,077 | 1,090,247 | 995.6 | 56.0 |
| Tver | 10,426 | 5,717 | 1,241,678 | 839.7 | 54.8 |
| Udmurtia | 10,344 | 5,691 | 1,460,624 | 708.2 | 55.0 |
| Kirov | 10,034 | 1,189 | 1,171,836 | 856.2 | 11.8 |
| Primorsky.Krai | 10,018 | 4,624 | 1,859,017 | 538.9 | 46.2 |
| Chuvashia | 9,683 | 4,418 | 1,194,876 | 810.4 | 45.6 |

| Region | Absolute Excess | Official COVID-19 Deaths | Population | Excess Rate | COVID-19 Share in Excess |
|---|---|---|---|---|---|
| Bryansk | 9,657 | 3,253 | 1,177,999 | 819.8 | 33.7 |
| Dagestan | 9,294 | 5,463 | 3,164,376 | 293.7 | 58.8 |
| Tambov | 9,153 | 4,337 | 992,766 | 922.0 | 47.4 |
| Khabarovsk | 8,567 | 5,690 | 1,298,362 | 659.8 | 66.4 |
| Kaluga | 7,814 | 4,651 | 1,059,372 | 737.6 | 59.5 |
| Arkhangelsk | 7,649 | 5,563 | 1,036,523 | 737.9 | 72.7 |
| Smolensk | 7,542 | 3,679 | 900,940 | 837.2 | 48.8 |
| Astrakhan | 7,300 | 3,937 | 968,950 | 753.4 | 53.9 |
| Vologda | 7,251 | 3,992 | 1,149,544 | 630.8 | 55.0 |
| Tomsk | 7,168 | 3,046 | 1,067,006 | 671.7 | 42.5 |
| Mordovia | 7,167 | 3,725 | 789,479 | 907.8 | 52.0 |
| Kurgan | 6,920 | 4,279 | 787,289 | 878.9 | 61.8 |
| Oryol | 6,421 | 4,409 | 720,550 | 891.1 | 68.7 |
| Kaliningrad | 6,342 | 3,932 | 1,022,401 | 620.4 | 62.0 |
| Ivanovo | 6,333 | 5,516 | 938,858 | 674.5 | 87.1 |
| Amur | 5,800 | 2,546 | 773,420 | 749.9 | 43.9 |
| Zabaykalsky.Krai | 5,534 | 3,228 | 1,013,773 | 545.9 | 58.3 |
| Mari.El | 5,421 | 1,254 | 679,852 | 797.4 | 23.1 |
| Karelia | 5,340 | 2,571 | 541,516 | 986.1 | 48.1 |
| Kostroma | 5,249 | 1,926 | 588,598 | 891.7 | 36.7 |
| Chechnya | 5,229 | 2,275 | 1,496,395 | 349.4 | 43.5 |
| Komi | 5,102 | 3,131 | 749,663 | 680.5 | 61.4 |
| Yakutia | 5,094 | 2,915 | 987,276 | 516.0 | 57.2 |
| Pskov | 5,064 | 1,970 | 604,900 | 837.1 | 38.9 |
| Murmansk | 4,942 | 3,206 | 678,672 | 728.2 | 64.9 |
| Novgorod | 4,788 | 2,560 | 587,758 | 814.6 | 53.5 |
| Buryatia | 4,626 | 2,931 | 980,500 | 471.8 | 63.4 |
| Kabardino.Balkaria | 4,145 | 1,997 | 900,734 | 460.2 | 48.2 |
| North.Ossetia | 4,011 | 2,108 | 690,494 | 580.9 | 52.5 |

| Region | Absolute Excess | Official COVID-19 Deaths | Population | Excess Rate | COVID-19 Share in Excess |
|---|---|---|---|---|---|
| Khakassia | 2,967 | 2,434 | 535,990 | 553.6 | 82.0 |
| Karachay.Cherkessia | 2,439 | 1,389 | 470,004 | 518.8 | 57.0 |
| Sakhalin | 2,375 | 543 | 469,560 | 505.7 | 22.9 |
| Adygea | 2,340 | 1,435 | 491,678 | 475.9 | 61.3 |
| Kalmykia | 1,566 | 1,147 | 268,600 | 583.2 | 73.2 |
| Kamchatka | 1,555 | 1,061 | 293,870 | 529.2 | 68.2 |
| Jewish.AO | 1,275 | 638 | 152,367 | 836.7 | 50.0 |
| Ingushetia | 1,070 | 996 | 504,012 | 212.3 | 93.1 |
| Altai | 1,025 | 700 | 211,114 | 485.6 | 68.3 |
| Tuva | 1,023 | 550 | 333,446 | 306.8 | 53.8 |
| Magadan | 738 | 363 | 137,319 | 537.1 | 49.2 |
| Chukotka | 138 | 44 | 47,916 | 287.4 | 32.0 |

Note: Regions are sorted in descending order according to the total absolute excess deaths count
